# Exercise prescription and strategies to promote the cross-education of strength: a scoping review

**DOI:** 10.1101/2022.09.12.22279860

**Authors:** Caleb C. Voskuil, Justin W. Andrushko, Boglarka S. Huddleston, Jonathan P. Farthing, Joshua C. Carr

## Abstract

**Objective:** To perform a scoping review of the literature on the experimental studies examining the role of resistance training frequency, intensity, the type of training, training volume, and adjuvant therapies on the cross-education of strength.

**Study Design:** Scoping Review.

**Literature Search:** The review was preregistered and performed with the search methodology described by the PRISMA extension for scoping reviews. CINAHL, MEDLINE, APA PsycInfo, SPORTDiscus, and Web of Science were systematically searched with grey literature searches and pearling of references thereafter.

**Study Selection Criteria:** Experiments were included in the review if they performed a unilateral resistance training intervention that directly compared the dose of a training variable on the cross-education response in healthy or clinical populations following a minimum of two weeks of training. Experiments must have reported maximal strength outcomes for the untrained limb.

**Data Synthesis:** For each experiment, the study population, intervention methods, the dosage of the training variable being studied, and the outcomes for the untrained, contralateral limb were identified and collectively synthesized.

**Results:** The search returned a total of 911 articles, 56 of which qualified for inclusion. The results show that experimental trials have been conducted on resistance training frequency (n = 4), intensity (n = 7), the type of training (n = 25), training volume (n = 3), and adjuvant therapies (n = 17) on the cross-education of strength.

**Conclusions:** This review synthesizes the available evidence regarding exercise design and prescription strategies to promote the cross-education of strength. It appears that traditional resistance training frequencies (ie., 2-3d/wk) at high intensities are effective at promoting cross-education. Eccentric muscle actions show additive benefits. There is experimental evidence that neuromodulatory techniques can augment cross-education when layered with unilateral resistance training versus training alone.

## Introduction

Cross-education has been known since the seminal work of Scripture et al^67^, but only recently has its clinical significance been given meaningful attention. The defining feature of cross-education is the transfer of muscle strength to the opposite (untrained) homologous limb following unilateral strength training. This adaptation has little relevance for individuals with well-functioning limbs. However, in a scenario where one limb is debilitated, unilateral training is a viable intervention to preserve neuromuscular function following orthopedic injury^31,45,56^ or neurotrauma.^17,51^ Despite evidence showing the therapeutic efficacy of cross-education, unilateral training is still poorly prescribed.^12^ This may be due, in part, to the lack of information available to clinicians to guide evidence-based decisions regarding the exercise prescription for cross-education.

The potential benefits of cross-education in rehabilitation settings cannot be understated. A common concern is that resistance training with only one limb will heighten the asymmetry between limbs in clinical contexts. However, the benefit of preserving muscle size and function likely outweighs the temporary asymmetry that could manifest with cross-education and can be corrected post-disuse or injury by shifting the focus of training to the opposite limb.^24^ In these scenarios, cross-education training provides a low-risk strategy that, at minimum, helps to prevent global deconditioning during periods of inactivity. The emerging evidence showing cross-education interventions preserve muscle strength and size during orthopedic limb immobilization^2,23,46,63,76^ offers more confidence that unilateral training has broad clinical applications. There are very little data from randomized clinical trials that have employed a cross-education intervention in patients. Of those that have, there is evidence^45,56,60^ showing cross-education provides superior adaptations compared to standard care, whereas others^83,84^ show similar outcomes with or without unilateral training. Differences in exercise design and prescription likely account for some of the disparity in these clinical trials.^56,60,83,84^

The transfer of strength to the opposite, untrained limb is mediated by neural adaptations. With that, exercise interventions that facilitate neural drive of descending motor commands may serve as promising strategies for enhancing the effect. Several reviews discuss the mechanisms and sites supporting cross-education.^1,14,25,32,50^ Given its neural basis, there is interest towards identifying optimal training prescriptions that maximize contralateral adaptations. A meta-analysis by Manca et al^48^ revealed that the type of muscle action used during unilateral training influences the magnitude of transfer, and Frazer et al^49^ outline adjuvant interventions showing promise for augmenting the transfer effect. A recent expert consensus statement^49^ provides direction for mechanistic inquiries into the cross-education effect as well as consensus-based recommendations for the design of unilateral training interventions aiming to promote strength transfer. Despite these advances, a thorough synthesis of the evidence regarding exercise design and prescription to optimize the cross-education of strength remains absent.

This scoping review addresses this gap by outlining the experiments that have examined the role of resistance training frequency, intensity, the type(s) of training, and training volume on the magnitude of cross-education. Emerging adjuvant therapies that may augment cross-education are also examined. Importantly, we focus on the cross-education of strength as it reflects the upper boundary of motor performance and is an accessible target across different clinics, conditions, and rehabilitation timelines.^30,42^ Our intention is to provide clinicians with evidence-based recommendations that optimize contralateral adaptations for individuals with asymmetrical limb function (i.e., orthopedic injury) who are unable to exercise both limbs.

## Methods

This scoping review adhered to the recommendations for the PRISMA extension for Scoping Reviews.^73^ The review topic was preregistered on Open Science Framework (osf.io/9sh5b) before study selection and data extraction. All literature searches were performed by an information scientist.

### Data Sources and Searches

A literature search protocol was developed through the identification of relevant health science databases available through the Texas Christian University Library and the construction of a Boolean search string. The following databases were systematically searched on December 20, 2021: CINAHL Complete, MEDLINE Complete, APA PsycInfo, SPORTDiscus with Full Text, and Web of Science. The following Boolean search string was utilized: ((cross education OR cross exercise OR cross transfer) OR (cross training NOT crossfit) OR (interlimb transfer OR “strength transfer”) OR bilateral transfer)) AND ((unilateral training OR unilateral exercise) OR (contralateral training OR contralateral exercise)). An expander was applied for equivalent subject terms. Results were limited to journal articles published after 1980 up to present time, and articles published in English. Three additional databases were searched for grey literature (ClinicalTrials.gov, HSRProj, and NIH RePORTER) and did not produce any additional results. Supplementary File Table 1 shows the search protocol executed in SPORTDiscus, including expanders, limiters, and number of items found.

### Study Selection

The initial search across the five databases yielded 2809 articles. Six additional articles that were known to the authors but did not show up in the database searches were added. A screen for duplicates removed 1904 articles and a set of 911 potential articles remained. The inclusion criteria utilized for this scoping review was designed to capture as many experiments as possible that have examined the role of resistance training frequency, training intensity, the type of training, training volume, and adjuvant interventions on the magnitude of cross-education. The following inclusion criteria was used for screening:

1. Participants: Investigations must have been performed on human participants; all ages, sexes, and abilities were included in the review.
2. Training comparisons: Studies must have included a direct comparison of a training variable and its influence on the magnitude of cross-education. The operational definitions of the training variables and their criteria for inclusion are composed of the following:
  a. Training Frequency: experiments must have compared the number of unilateral resistance training sessions within a defined period.
  b. Training Intensity: experiments must have compared the load and/or resistance relative to maximal strength levels for a unilateral resistance exercise.
  c. Type of Training: experiments must have compared the type of muscle action, joint action, or the limb involved within the unilateral resistance training intervention.
  d. Volume of Training: experiments must have compared the amount or structure of the sets and/or repetitions of unilateral resistance training within the intervention.
  e. Adjuvant Intervention: experiments must have compared the effects of a treatment intervention with unilateral training compared to unilateral training alone on the magnitude of cross-education.
3. Since this review focuses on the dose-response properties of resistance training variable prescription on the magnitude of cross-education, a minimum of two weeks of training must have been performed.
4. Outcome measures: Studies must have reported measurements of maximal strength to quantify the magnitude of cross-education.
5. Additional considerations: Review articles and conference abstracts were not included in this review.

## Results

715 studies were excluded based on title, keywords, and abstract screening, and 156 were excluded based on full-text screening. A reference check was conducted on the remaining 40 articles, which added 16 new articles. A total of 56 articles met inclusion criteria and were retained for inclusion in this scoping review. Figure 1 provides an overview of the study selection process.

**Figure 1.**
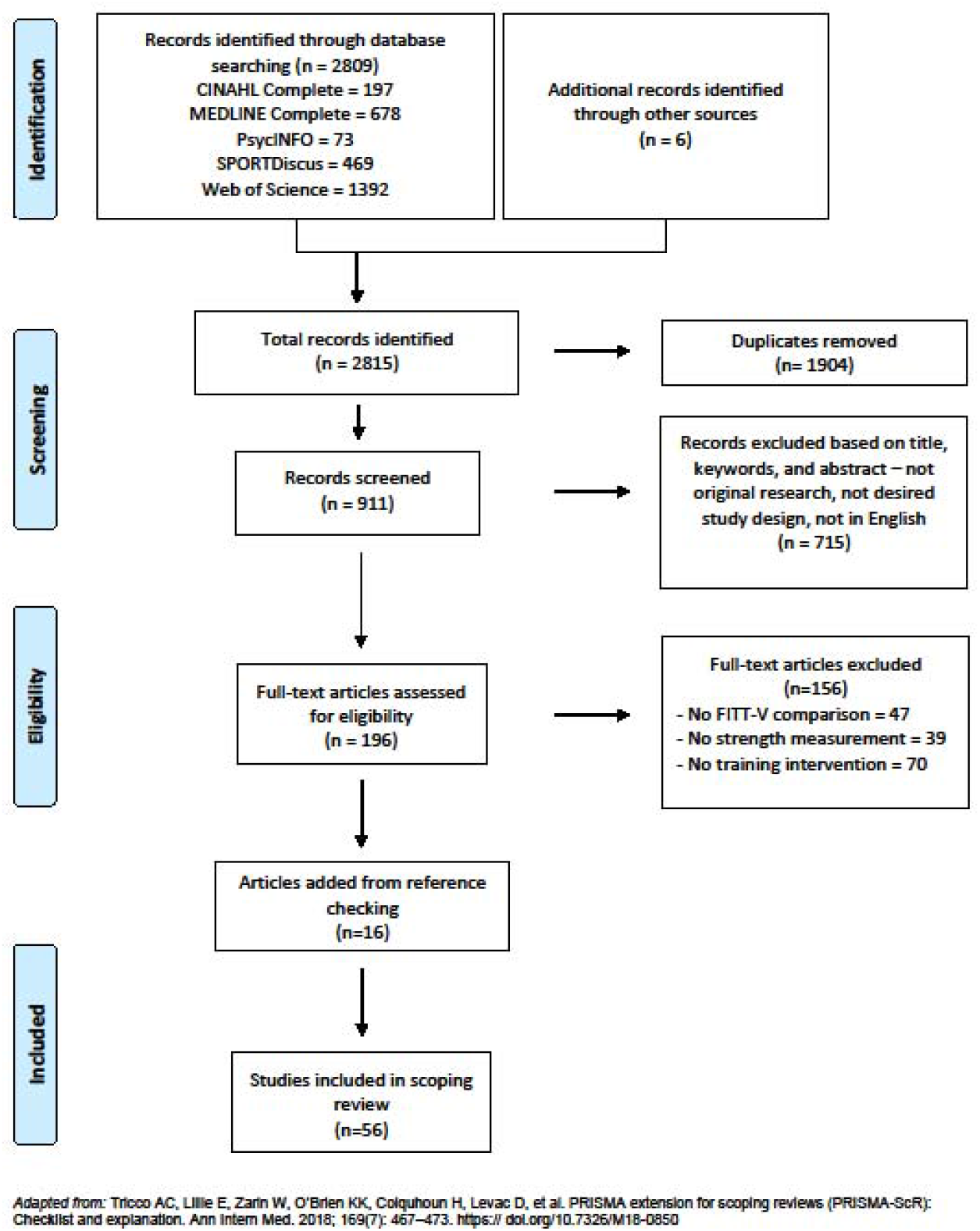
PRISMA Flowchart of Article Search and Selection

## Study characteristics

Of the articles included in the review, the total sample size across all 56 articles was N = 1801 with a median of n = 31 per experiment. The median number of groups per experiment was n = 3. There was considerable range in the sample size per experiment (n = 9-115). A total of six studies compared exercise prescription variables in clinical populations and one during orthopedic immobilization. Most studies (n=49) were performed in younger adults (median age: 23 years), while four articles were performed in adolescents (median age: 11.5 years) and three performed with older adults (median age: 62 years). There were three studies that did not report the sex of the sample; of those that did, 66% were male and 34% were female.

There were three articles comparing more than one exercise prescription variable.^13,52,58^ Of these, the primary aim of the respective experiment was used for categorization within the review, with secondary aims incorporated when necessary. Four articles met inclusion criteria but were not designed to examine cross-education, but as a control measure during a unilateral intervention.

## Discussion

The American College of Sports Medicine provides general recommendations for resistance training design and prescription for the major resistance training variables: Frequency, Intensity, Type, and Volume.^43^ These variables are manipulated within a resistance training session to provoke specific adaptations (i.e., strength, hypertrophy, endurance), yet there are no standardized recommendations for unilateral training to promote the cross-education of strength. By synthesizing the available data comparing the exercise design and prescription variables for unilateral training in this scoping review, we map the current knowledge on this topic and outline a general direction for design strategies to support the cross-education of strength. We summarize the findings for each training variable below and outline general recommendations in Table 3.

**Table 3.** General recommendations for unilateral resistance training to promote the cross- education of strength.

| Variable | Recommendation | Evidence |
| --- | --- | --- |
| **Frequency** | Unilateral exercise should be performed 2-3 days per week.<br>Increasing the training frequency may increase rate of cross-education. | 3, 9, 60, 61 |
| **Intensity** | To promote cross-education, unilateral exercise should be performed at relatively high intensities of effort. | 5, 10, 13, 57, 64 |
| **Type** | Multi-joint and single joint exercise can promote cross-education<br>Cross-education manifests for the upper, lower, dominant, and non-dominant limbs.<br>Isokinetic, isotonic, and isometric exercise all promote cross-education.<br>Eccentric muscle actions should be incorporated and at times emphasized during unilateral exercise. | 6, 7, 15, 16, 21, 22, 29, 31, 34-36, 40, 47, 52, 53, 62, 65, 66, 68, 69, 74, 76-78 |
| **Volume** | Multiple sets with progressive increases in volume are recommended to promote cross-education. | 13,19,58 |

### Frequency

The role of training frequency on the magnitude of cross-education has been examined with handgrip training^3,9^ and knee extension training following anterior cruciate ligament reconstruction (ACLR).^60,61^ The grip training experiments compare a traditional frequency of training (ie., 3×/wk) with a high frequency of training at the same intensity (i.e., 7×/wk or 10×/wk respectively). The data shows that when handgrip training is volume-matched, high-frequency training does not further the magnitude of cross-education compared to a traditional training frequency.^3,9^ However, with a unique study design, Barss et al^3^ show that high-frequency training (daily) increases the rate at which cross-education manifests. Specifically, cross-education was observed in the daily training group following the 15^th^ session (∼2 weeks) of the training intervention, while cross-education was shown for the traditional training group after completion of the 12^th^ session (∼ 1 month). Similar findings are shown in experiments examining the role of training frequency during the early phase of rehabilitation following ACLR.^60,61^ By comparing different frequencies (3d/wk versus 5d/wk) of unilateral knee extension training to standard care, the authors show that strength outcomes are improved in the reconstructed knee of the training groups more than standard care control group following 8 weeks of eccentric-based unilateral training (5 sets × 6 reps; 80% eccentric 1RM). However, there were no significant differences in the outcomes for the reconstructed knee between the two experimental groups.

More specifically, the recovery of isometric knee extension strength to pre-operative levels was lowest in the standard treatment group with ∼57% recovery versus ∼80% and ∼77% strength recovery for the 3×/wk and 5×/wk groups, respectively. The available data shows that increasing training frequency does not further the magnitude of cross-education.^3,9,60,61^ However, high frequency isometric training increases the rate at which cross-education manifests, showing similar strength improvements in ∼half the time.^3^ Overall, these findings indicate that increasing the training frequency influences the rate of cross-education but not the magnitude.

### Intensity

The intensity of resistance training is arguably the most important variable in the exercise prescription and is a key moderator of the neural adaptations that manifest from training.^27^ The role of training intensity on cross-education has been examined through submaximal handgrip training,^70,79^ elbow flexion,^57,64^ unilateral leg press in adolescents,^5,10^ and knee extension.^13^ The comparison of training loads (50%, 33%, and 30%, of maximal strength) prescribed to submaximal handgrip endurance training (2 sets to failure; 5-6×/wk for 6 wks) demonstrates that higher intensity loads exhibit greater cross-education than lower intensities. This positive relationship between the training intensity and the magnitude of cross-education is supported during both volume-matched and non-volume-matched elbow flexion training.^57,64^ During higher intensity training (≥ 80% 1RM) a greater magnitude of cross-education is observed when compared to lower intensity loads (≤ 40%).^57,64^ Additionally, training at higher intensities may increase the rate of cross-education, showing intensity-dependent outcomes following a single week of training.^64^ Training intensity comparisons during lower-body training show similar results.^5,10,13^ In adolescents performing volume-matched high intensity (5RM) or low intensity (20RM) unilateral leg press training, high-intensity training demonstrates ∼25% and 20% greater contralateral strength increases in knee extension and flexion, respectively, compared to low-intensity.^5,10^ Similar findings are observed during knee extension in non-clinical adults.^13^ When training is performed at 75% 1RM for a defined volume (6 sets × 5 reps) or until failure, the magnitude of cross-education is greater than training at 25% 1RM to failure.^13^ The available data consistently demonstrates superior cross-education effects when unilateral training is performed at higher versus lower intensities.^5,10,13,57,64,70,79^

### Type

#### Muscle action

Several experiments have examined how the type of muscle action used during unilateral training affects the magnitude of cross-education. Specifically, concentric versus eccentric training has been compared during isokinetic knee extensions,^31,35,36,68,69^ isotonic knee extensions,^77,78^ elbow flexion,^34,65,74,76^ and isokinetic wrist flexion.^40^ Most experiments show eccentric training results in superior cross-education versus concentric training and provides more robust transfer effects to other muscle actions.^34,35,36,40,68,69,76^ Even in the experiments showing similar magnitudes of cross-education, it seems that cross-education is better preserved during detraining following eccentric versus concentric training.^65,77,78^ The unique influence of unilateral eccentric training has been examined in clinically relevant scenarios.^31,76^ Compared to traditional isotonic training with both muscle actions (ie., concentric-eccentric), eccentric training shows greater strength preservation effects following four weeks of arm immobilization for the immobilized arm.^76^ Eccentric versus concentric training of the non-affected knee extensors in ACLR patients shows that both types of unilateral training resulted in greater strength recovery of the reconstructed knee compared to standard care, but superior effect sizes were shown for the eccentric versus concentric training group at 12- and 24-weeks post- surgery.^31^ The experiments captured by our search show that unilateral eccentric training promotes more robust cross-education effects than concentric training in non-clinical and clinical populations.

#### Joint action

Experiments have examined how the type of joint action (i.e., isometric, isokinetic, isotonic) used during unilateral training influences the cross-education response^16,47,62^ and some also compare the training velocities (i.e., slower versus faster) during single-joint^21,58^ and multi-joint exercise.^52,53^ Interestingly, available evidence utilizing single leg squats,^53^ isotonic^58^ and isokinetic elbow flexion^21^ suggest training velocity influences cross-education. Unilateral multi- joint training at lower training velocities (duration of eccentric phase: 6-sec versus 3-sec) shows a greater magnitude of cross-education.^53^ Conversely, when training velocity is low during isokinetic or isotonic training of the elbow flexors, there is less transfer to the contralateral limb compared to training at higher velocities. However, when training velocity, volume, and intensity of training are ∼matched, there is no difference between isokinetic versus isotonic resistance training on the magnitude of cross-education.^16^ Additional comparisons of joint action type on cross-education are challenging due to differences in training intensities. Specifically, maximal isometric knee extensions promote cross-education following three months of unilateral training whereas no cross-education was shown following low-intensity (6.4 kg) isotonic knee extensions in Antarctic explorers.^62^ Similar findings are shown in patients with osteoarthritis who were separated into isometric, isokinetic, and isotonic training groups and performed unilateral training 5×/wk for 3wks.^47^ Although the repetition volume was matched for each training session between groups, the isometric training was maximal, the isotonic training was low-intensity and fixed (1.5 kg), and the isokinetic training was concentric-only.^47^ After the intervention, only the isometric group showed significant improvements in contralateral strength.^47^ These results^47,62^ emphasize the importance of unilateral training intensity to promote the cross-education of strength. When the intensity of isotonic training is matched, however, it seems that training at an extended (0° - 50°) versus flexed (80° - 130°) joint angle and range of motion promotes greater cross-education.^66^ These studies^16,21,47,52,53,62,66^ collectively show that isometric, isokinetic, and isotonic joint actions will promote the cross-education of strength following unilateral training of sufficient intensity. The implications being that regardless of the equipment availability within a clinic, favorable contralateral adaptations may be achieved.

#### Limb

In addition to the type of muscle and joint action, the magnitude of cross-education between the upper versus lower limb^7,29^ as well as the direction of transfer between dominant and non- dominant limbs^6,15,22,75^ have been examined. The magnitude of cross-education is similar between the upper and lower body^7,29^ and presents in both the dominant and non-dominant limbs.^6,15,22,75^ As the chance of sustaining an injury is similar across limbs, these findings offer support for the broad implementation of unilateral training when indicated.

### Volume

The volume of resistance training^13,58^ and set configuration^19^ have been examined for their effects on cross-education. When comparing training volumes during unilateral knee extension^13,59^ and elbow flexion,^19,58^ a dose-response relationship emerges between unilateral training volume and the magnitude of cross-education. However, it appears there is a threshold as well as diminishing returns for this relationship. For instance, no contralateral improvements are shown following unilateral knee extension training 1×wk every other week^59^ or following 30 discrete repetitions of elbow flexion 1×wk for 5 weeks.^19^ Cross-education is observed for the elbow flexors following a single set and three sets at a 6-8 RM load 3×/wk for 6 weeks, but greater transfer occurs when the volume of training increases from a single set to three sets.^58^ There is no additional contralateral benefit, however, when high-intensity (75% 1RM) training is performed to failure (6 sets × failure) versus a defined volume (6 sets × 5 reps).^13^ These studies suggest traditional strength training volume configurations (i.e., multiple sets of < 8 repetitions) at high intensities produces the most robust cross-education effect.^13,19,58,59^

### Adjuvant interventions

There has been considerable interest in augmenting the magnitude of cross-education with various adjuvant interventions. This likely reflects the broad clinical interest in understanding and applying cross-education as a neurorehabilitation technique. The brief sections below are organized in the domains of *Stimulation, Mirror Training & Mental Imagery, Blood Flow Restriction*, and a collection of unique approaches.

#### Stimulation

The use of muscle,^8,37,39^ nerve,^4^ and brain^33^ stimulation techniques in conjunction with unilateral training versus training alone show unique and somewhat conflicting results for the untrained limb. The studies investigating muscle stimulation have all done so for the knee extensors and show favorable^8,37^ or similar^39,81^ results on the magnitude of cross-education versus training alone. Those showing additive benefits of unilateral training with muscle stimulation used high training intensities^8^ and eccentric contractions,^37^ while the interventions showing similar outcomes used lower intensities of isometric training.^39,81^ One study^33^ shows that applying anodal transcranial direct current stimulation (tDCS) over the ipsilateral motor cortex (the ‘untrained’ M1) during unilateral training promotes a slightly greater magnitude of cross-education (∼12.5% versus 9.4%) and retention (13% versus 7.6%) following a brief period of detraining versus resistance training alone for the elbow flexors. In contrast, applying transcutaneous nerve stimulation to the radial nerve of the trained limb concurrently with training impairs the magnitude of cross-education compared to training alone for the wrist extensors.^4^ These studies collectively show that stimulation techniques can modify the magnitude of cross-education following unilateral training versus training alone. The use of neuromuscular electrical stimulation (NMES), tDCS, and other stimulation techniques^81^ seem to be promising adjuvant interventions to layer within unilateral training prescription, though the exact prescription for these techniques requires more investigation (see Zhou et al^82^ for more on this topic).

#### Mirror Training & Mental Imagery

Illusionary mirror training as well as mental imagery training are believed to activate the mirror neuron system^38,85^ and both forms of training provide an accessible avenue for patient-driven rehabilitation. The mirror training hypothesis^38^ has been investigated in neurologically intact individuals^86^ and in individuals living with stroke.^18,71^ These three studies compare the magnitude of cross-education while performing unilateral resistance training with and without illusionary mirror visual feedback during wrist flexion,^86^ elbow extension,^18^ and dorsiflexion.^71^ The data from neurologically intact individuals show ∼27% greater magnitude of cross-education in the mirror training group compared to the group that did not receive mirror visual feedback during unilateral training.^86^ The data from individuals living with stroke shows that when training the less affected limb with or without a mirror illusion, the mirror group displays cross-education of elbow extensor strength (∼15%), but no cross-education was observed following training without a mirror (-0.2%). Using the same training intervention as Ehrensberger et al,^18^ dorsiflexion training^71^ of the less affected limb with a mirror illusion shows a moderate, non-significant improvement in the contralateral dorsiflexors strength (∼6%, ES = 0.40, *p* = 0.160) but less of an effect when training is performed without a mirror (-0.1%, ES < 0.00, *p* = 0.956). It is important to note the large, non-significant between-group effect for the elbow extensors (*p* = 0.056, ES = 0.70) and negligible between-group effect for the dorsiflexors (*p* = 0.956, ES = 0.10) in the experiments studying individuals living with stroke. The two studies comparing mental imagery training versus unilateral training alone on the magnitude of cross-education report contrasting findings.^20,80^ Cross-education was similar between mental imagery versus the unilateral training group for the small muscles of the hand,^80^ whereas only the unilateral training group demonstrated cross-education for a novel grip task.^20^ The evidence shows that mirror training seems to be a promising adjuvant for cross-education in neurologically intact populations and individuals living with stroke for tasks involving the upper limb, but it may be that isotonic versus isometric tasks provide a greater training stimulus. The evidence does not support the notion that mental imagery provides additive benefits over unilateral training alone, however, the limited comparisons open the door for novel experimental designs to examine this question further.

#### Blood Flow Restriction

There have been three experiments comparing the effect of unilateral training with versus without blood flow restriction (BFR) to the training limb. The data from the elbow flexors shows that following 20 training sessions of 3 sets × 10 repetitions at 50% 1RM, there was either no cross-education^44^ or a significant level of strength improvement for the untrained arm that was not enhanced by BFR.^54^ Similar findings are shown when comparing the influence of high-intensity (75%1RM) resistance training versus low-intensity (20%1RM) BFR training of the plantar flexors. Following 20 training sessions, no cross-education of strength and no benefit of BFR was observed.^55^ The available data indicates that adding BFR to a unilateral training intervention provides no benefit for the contralateral, untrained limb.

#### Whole Body Vibration, Protein Supplementation, and Hypoxia

The use of unilateral resistance training with whole body vibration,^28^ protein supplementation,^11^ and training under hypoxic conditions^41^ have been examined. Performing high-intensity unilateral split squats with or without the rear leg on a vibrating platform drastically increases split squat 1RM for the contralateral leg,^28^ but vibration (+52%, ES = 0.98) does not provide significantly greater benefits versus training without vibration (+35%, ES = 0.99). Interestingly, in the protein supplement study, participants who consumed 20 g of whey protein and 6.2 g of leucine before and after high-intensity training of the knee extensors for 8 weeks showed significantly greater magnitudes of cross-education (∼15%) compared to the placebo (2.8%) and control (4.6%) groups.^11^ Lastly, training under hypoxic conditions, which elevated growth hormone while resistance training, offered no significant benefit towards the magnitude of cross-education (40%) compared to normoxic conditions (24%).^41^

### Limitations

Our focus on the cross-education of strength neglects the cross-education of skill that is well known and clinically relevant. The quality of the included studies varied considerably as some studies use a parallel group design with, and others without, a non-interventional control group. The participants who underwent training were mostly free of injury or disease, which may limit the generalizability of these findings for the intended patient populations. The relevance of adhering to reporting standards for exercise interventions is obvious^72^ and needed for the replication, development, and clinical translation of unilateral resistance training paradigms. This is a critical point for cross-education interventions as principles of motor learning (i.e., instruction, feedback, attentional focus) influence skill acquisition.

### Clinical Implications

Unilateral resistance training provides a low-cost, accessible rehabilitation strategy for individuals unable to exercise one limb due to injury or neurotrauma. Clinicians can use this review as a road map for developing unilateral rehabilitation interventions that promote favorable outcomes for the affected limb in patients with asymmetrical limb function. These practices can be integrated within multiple levels of athlete-patient care.

### Conclusions

This review maps the available evidence regarding the dose-response properties of unilateral resistance training frequency, intensity, the type(s) of training, and training volume on the cross- education of strength and outlines adjuvant interventions that may augment the transfer effects. The evidence shows that exercise design and prescription moderate the cross-education of strength following short-term training interventions in clinical and non-clinical populations. The key findings indicate that training 2-3×wk at relatively high intensities of effort are optimal for training. Whether training is single-joint or multi-joint, emphasizing eccentric muscle actions appears beneficial. As training progresses, so should the volume of unilateral exercise. Clinicians may consider using these recommendations as a starting point for implementing cross-education as a rehabilitation strategy. These recommendations are accessible, low-risk, and provide the patient with autonomy regarding their rehabilitation.

### Key points

#### Findings

The cross-education of strength is moderated by exercise design and prescription in clinical and non-clinical populations.

#### Implications

This review synthesizes the available evidence regarding exercise design strategies for unilateral resistance training and provides evidence-based recommendations for the prescription of unilateral training to maximize the cross-education of strength.

#### Caution

Unilateral training is another tool in a clinician’s toolbox, not a panacea, the training should align with the needs of the patient and the objectives of the rehabilitative phase.

## Supporting information

Supplementary File: Table 1. SPORTDiscus Search

Supplementary File: Table 2.Table of Article Characteristics

Supplementary File 3: PRISMA ScR Checklist

## Data Availability

All data produced in the present work are contained in the manuscript.

