## Supplementary File: Table 1. SPORTDiscus Search for "Exercise prescription and strategies to promote the cross-education of strength: a scoping review"

EBSCOhost

Monday, December 20, 2021 6:23:37 PM

| # | Query | Limiters/Expanders | Last Run Via | Results |
|---|---|---|---|---|
| S11 | S7 AND S8 | Limiters - Published Date: 19800101-20211231<br>Expanders - Apply equivalent subjects<br>Narrow by Language: - english<br>Search modes - Find all my search terms | Interface - EBSCOhost Research Databases<br>Search Screen - Advanced Search<br>Database - SPORTDiscus with Full Text | 469 |
| S10 | S7 AND S8 | Limiters - Published Date: 19800101-20211231<br>Expanders - Apply equivalent subjects<br>Search modes - Find all my search terms | Interface - EBSCOhost Research Databases<br>Search Screen - Advanced Search<br>Database - SPORTDiscus with Full Text | 481 |
| S9 | S7 AND S8 | Expanders - Apply equivalent subjects<br>Search modes - Find all my search terms | Interface - EBSCOhost Research Databases<br>Search Screen - Advanced Search<br>Database - SPORTDiscus with Full Text | 486 |
| S8 | S4 OR S5 | Expanders - Apply equivalent subjects<br>Search modes - Find all my search terms | Interface - EBSCOhost Research Databases<br>Search Screen - Advanced Search<br>Database - SPORTDiscus with Full Text | 3,349 |
| S7 | S1 OR S2 OR S3 OR S6 | Expanders - Apply equivalent subjects<br>Search modes - Find all my search terms | Interface - EBSCOhost Research Databases<br>Search Screen - Advanced Search<br>Database - SPORTDiscus with Full Text | 24,610 |
| S6 | bilateral transfer | Expanders - Apply equivalent subjects | Interface - EBSCOhost Research Databases<br>Search Screen - Advanced | 179 |

| | | | | |
|---|---|---|---|---|
| | | Search modes - Find all my search terms | Search<br>Database - SPORTDiscus with Full Text | |
| S5 | contralateral training OR contralateral exercise | Expanders - Apply equivalent subjects<br>Search modes - Find all my search terms | Interface - EBSCOhost Research Databases<br>Search Screen - Advanced Search<br>Database - SPORTDiscus with Full Text | 1,042 |
| S4 | unilateral training OR unilateral exercise | Expanders - Apply equivalent subjects<br>Search modes - Find all my search terms | Interface - EBSCOhost Research Databases<br>Search Screen - Advanced Search<br>Database - SPORTDiscus with Full Text | 2,449 |
| S3 | interlimb transfer OR "strength transfer" | Expanders - Apply equivalent subjects<br>Search modes - Find all my search terms | Interface - EBSCOhost Research Databases<br>Search Screen - Advanced Search<br>Database - SPORTDiscus with Full Text | 47 |
| S2 | cross training NOT crossfit | Expanders - Apply equivalent subjects<br>Search modes - Find all my search terms | Interface - EBSCOhost Research Databases<br>Search Screen - Advanced Search<br>Database - SPORTDiscus with Full Text | 9,537 |
| S1 | cross education OR cross exercise OR cross transfer | Expanders - Apply equivalent subjects<br>Search modes - Find all my search terms | Interface - EBSCOhost Research Databases<br>Search Screen - Advanced Search<br>Database - SPORTDiscus with Full Text | 18,480 |
